# Association between triglyceride glucose index and heart failure with preserved ejection fraction in patients with coronary heart disease — a RCSCD-TCM study in China

**DOI:** 10.1101/2023.10.31.23297884

**Authors:** Zhu Li, Xiang Fan, Yijia Liu, Lu Yu, Yuanyuan He, Lin Li, Shan Gao, Wei Chen, Rongrong Yang, Chunquan Yu

## Abstract

**Background:** The triglyceride glucose (TyG) index serves as a surrogate indicator of insulin resistance. However, there is limited evidence on the association between the TyG index and heart failure with preserved ejection fraction (HFpEF) in patients with coronary heart disease (CHD).

**Methods:** The 62,794 CHD patients were included used to analyze the relationship between the TyG index and heart failure (HF) in CHD patients. Of these, 8,606 patients who underwent echocardiography were included to identify different types of HF, including HF with reduced ejection fraction (HFrEF), HF with intermediate-range ejection fraction (HFmrEF), and HFpEF. Logistic regression was used to analyze the relationship between the TyG index and HFpEF in CHD patients. The relationship between the TyG index and HFpEF according to sex, age, blood lipids and blood pressure states were also assessed.

**Results:** A baseline analysis of CHD patients divided into four groups according to the tertile level of the TyG index showed that there were significant differences in related parameters between the groups. In the multi-adjusted modles, the TyG index was significantly associated with the risk of HFpEF (OR: 1.56; 95% CI: 1.08–1.23). In addition, the TyG index of CHD patients was significantly associated with HFpEF in elderly (> 60 years old) patients (OR:1.19; 95% CI: 1.10-1.48), hypertension (OR:1.17; 95% CI: 1.10-1.25) and dyslipidemia (OR:1.16; 95% CI: 1.08-1.23). The association between the TyG index and HFpEF was not affected by sex. And the association between the TyG index of female and HFpEF was (OR:1.21; 95% CI: 1.10-1.34), which was higher than that of male (OR:1.11; 95% CI: 1.02-1.21).

**Conclusions:** This study demonstrated a significant association of the TyG index and HFpEF in CHD patients. In this study, the results show that the TyG index was independently associated with HFpEF in hypertension, dyslipidemia, and elder patients (> 60 years old). In addition, the association between the TyG index and HFpEF in CHD patients was higher in female.

Graphical abstract

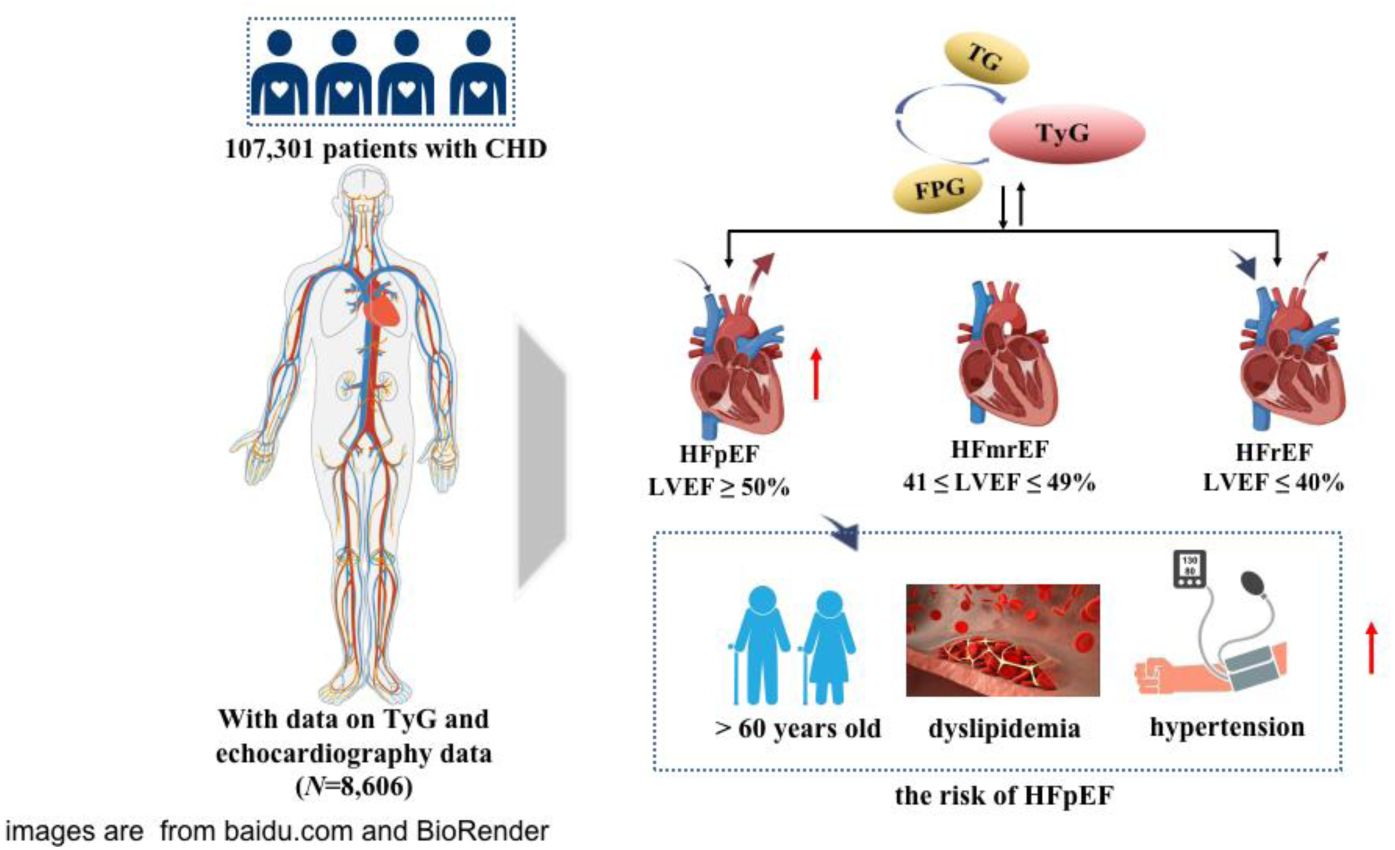

## Background

Coronary heart disease (CHD) is a kind of coronary atherosclerotic heart disease. Its pathological process leads to coronary artery stenosis, which leads to myocardial ischemia, myocardial necrosis, myocardial systolic dysfunction, and heart failure (HF) due to decreased ejection capacity [1–2]. HF with preserved ejection fraction (HFpEF) is a common type of HF, and about 50% of HF patients suffer from HFpEF [3]. According to the guideline of HF [4], the percentage of hospitalization of HFpEF in total HF patients is increasing year by year. It is predicted that by 2020, 65% of hospitalized patients with HF will have left ventricle ejection fraction (LVEF) >40%. Generally speaking, HFpEF has a high morbidity, readmission rate and readmission mortality, and HFpEF and HF with reduced ejection fraction (HFrEF) have similar morbidity severity and mortality. Therefore, the prevention and treatment of this disease need further research and discussion.

The triglyceride glucose (TyG) is used as a marker of insulin resistance (IR), leading to the occurrence of NCD [5–6]. Many studies have shown that the TyG index is related to the high prevalence of CAD and the increased risk of major adverse cardiovascular and cerebrovascular events [7–11]. Previous study showed that the TyG index and CAP showed a significant association in CHD patients [12]. A Mendelian Randomization Study showed that the TyG index can be used as a more sensitive prediagnostic indicator of CVD, and that could provide quantitative risk for cardiometabolic outcomes including HF [13]. Recently studyshowed that the TyG index is a novel biomarker of myocardial fibrosis in HF patients and can be considered as a useful risk stratification metric in the management of HF [14]. However, there are no relevant studies to investigate the relationship between the TyG index and HF and the different typing of HF in CHD patients, especially HFpEF.

Therefore, the purpose of this study was to clarify the association of the TyG index and HF in CHD patients, and to further investigate the association of the TyG index and the different typing of HF, especially HFpEF. It is hoped that in the clinical treatment of CHD, through the identification of simpler biochemical indicators to prevent the risk of HF.

## Methods

### Patients

This study is a large-scale, multi-center retrospective study. The participants included 107,301 CHD inpatients who were admitted to 6 hospitals in Tianjin from January 1, 2014 to September 30, 2020. The root investigation study design excluded age less than 35years or older than 80 years, patients with tumor, infectious, or severe liver or kidney disease, and patients lacking data on TG, FPG, and echocardiography data. A total of 8,606 patients who underwent echocardiography were included to identify different types of HF, including HFrEF, HF with intermediate-range ejection fraction (HFmrEF), and HFpEF. A flowchart of the patients recruitment was shown in Figure 1. This study was approved by the ethics committee of the Tianjin University of Traditional Chinese Medicine (TJUTCM-EC20190008) and registered with the Chinese Clinical Trial Registry (ChiCTR-1900024535) and in ClinicalTrials.gov (NCT04026724).

**Figure 1.**
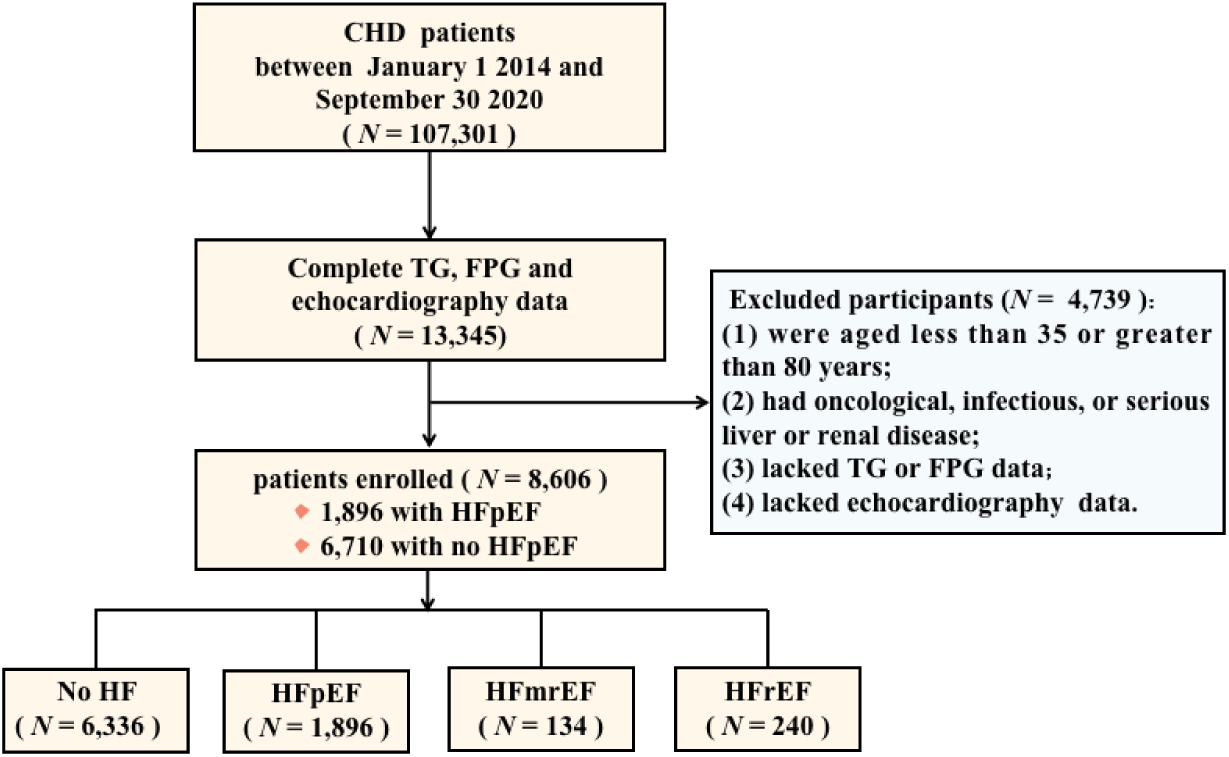
Flow chart of patient recruitment.

### Data collection

In this study, age, sex, smoking, drinking, and medication history of patients were recorded through standard structured questionnaires [15,16]. All participants collected fasting venous blood samples in early morning. FPG, total cholesterol (TC), high-density lipoprotein cholesterol (HDL-C), and TG, low-density lipoprotein cholesterol (LDL-C), glycated haemoglobin (HbA1c) were measured on automatic haematology analyser. And strictly follow the laboratory standard procedures for quality control. The TyG index was calculated as Ln[TG (mg/dL) × FPG (mg/dL)/2] [17]. Hyperlipidemia was defined as TC ≥ 6.2mmol/L (240mg/dL), TG ≥ 2.3mmol/L (200mg/dL), LDL-C ≥ 4.1mmol/L (160mg/dL), or HDL-C ≤ 1.0mmol/L (40mg/dL) [18,19]. Systolic blood pressure (SBP) and diastolic blood pressure (DBP) were measured by experienced technicians at the heart level using automatic blood pressure monitors. SBP ≥ 130 mmHg or a DBP ≥ 80 mmHg was defined as hypertension [20]. Normal glucose tolerance (NGT) was defined as FPG <5.6 mmol/L or HbA1c <5.7%, Pre-DM was defined as FPG 5.6-6.9 mmol/L or HbA1c 5.7-6.4%, DM was defined as FPG ≥ 7.0 mmol/L or HbA1c ≥ 6.5% [21].

The HF diagnosis was recorded in the data base of RCSCD-TCM. HF including congestive HF, left ventricular failure, New York Heart Association (NYHA) Heart Function class Ⅱ-IV [22], and unspecified HF. Different typing of HF use left ventricle ejection fraction (LVEF) measured using echocardiography as a cut-off for inclusion/exclusion criteria. According to European Society of Cardiology (ESC) Guidelines [23], HF patients into 3 groups/categories based on LVEF: HFpEF patients considered those with LVEF ≥ 50%. HFmrEF patients considered those with 41 ≤ LVEF ≤ 49%. HFrEF patients considered those with LVEF ≤ 40%.

### Statistical analyses

The characteristics of participants in the different groups were compared using *χ*^2^ tests for categorical variables, Mann-Whitney U test and Kruskal-Wallis H test for continuous variables. The odds ratios (ORs) and 95% confidence intervals (CIs) of CAP were estimated for the TyG index using logistic regressions. Age, sex, SBP, DBP, HbA1c, TC, HDL-C, LDL-C, smoking, drinking, hypertension, hyperlipidemia, use of antihypertensives, use of antilipidemic were considered potential confounders in this association. The collinearity of different models was tested before logstic regressions. Missing values are imputed by multiple imputation method. All statistical analyses were performed using SPSS 24.0 (IBM Corp, New York, NY, USA).

## Results

### Baseline characteristics

Of these, 8,606 patients underwent echocardiography, among them, 1896 patients had HFpEF. The average age of the participants was 65 years old, and the proportion of male (56.8%) was higher than that of female (43.2%) **(Adittional file Table S1)**. The subjects were divided into three groups according to the tertile level of the TyG index (T1: TyG index < 10.09, T2: 10.09 ≤ TyG index ≤ 10.71, T3: TyG index > 10.71). Generally speaking, FPG, TC, TG, LDL-C, HbA1c, the proportion of HFrEF, HFmrEF, HFpEF, hyperlipidemia, use of antihypertensives, and use of antilipidemic were positively associated with the tertile level of the TyG index, while HDL-C, the proportion of smoking and drinking were negatively associated with the tertile level of the TyG index (**Table 1).**

**Table 1.**
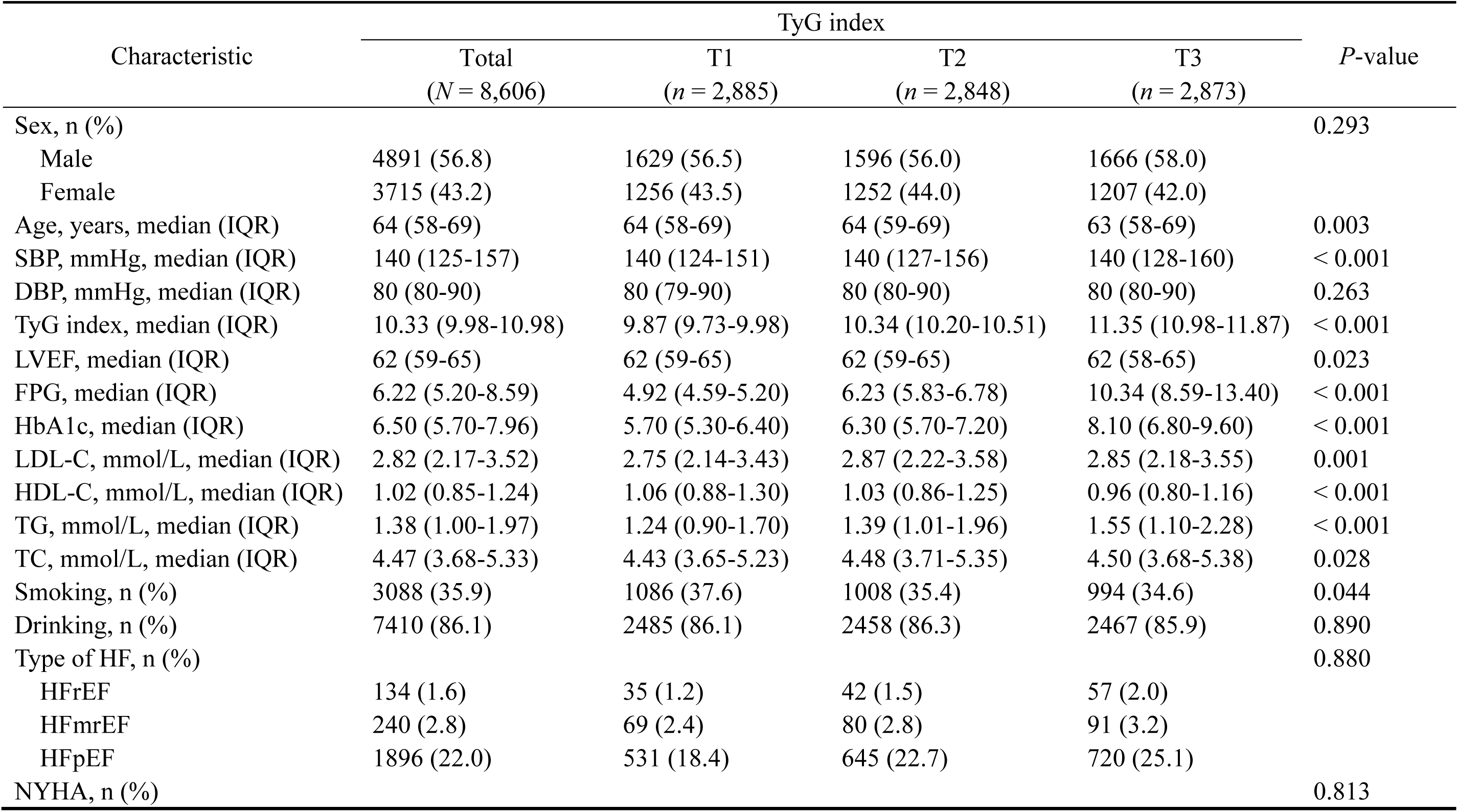
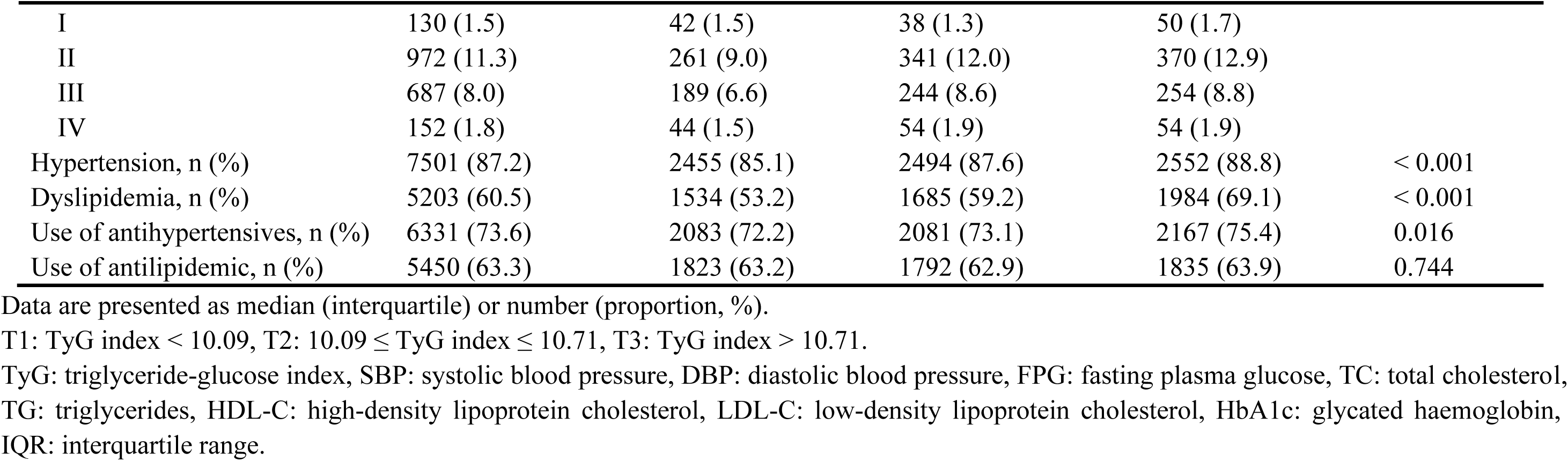
General characteristics of the study participants according to the TyG index.

### Association between the TyG index and the risk of HFpEF

As shown in **Table 2**, logistic regression models were constructed to show that the TyG index was significantly related to HFpEF before and after multivariate adjustment (*P* < 0.001). When the TyG index was analyzed as a continuous variable, it was significantly associated with HFpEF (OR: 1.56; 95% CI: 1.08–1.23). When the TyG index served as a classified variable, the risk of HFpEF for patients in T3 was 1.47 times greater than the risk for patients in T1. The association of the TyG index and the risk of different types of HF were further evaluated, including HFrEF, HFmrEF, and HFpEF. The results show that the association remained signifcantly different **(Adittional file Table S2)**.

**Table 2.**
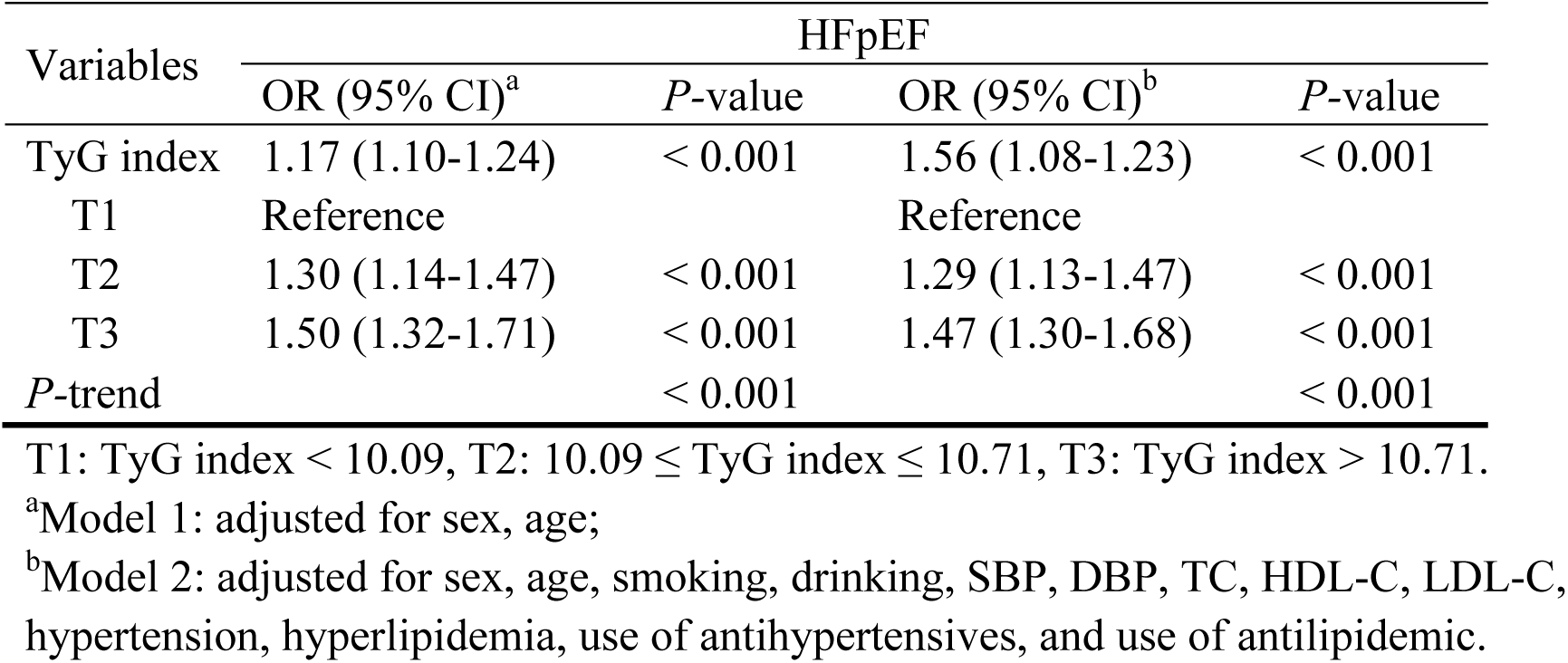
Association between the TyG index and the risk of HFpEF.

### Association between TyG index and the risk of carotid artery plaques according to sex and age

Association between the TyG index and the risk of HFpEF according to age and sex were shown in **Table 3**. After multivariate adjustment, the TyG index of CHD patients was significantly associated with HFpEF in elderly (> 60 years old) patients (OR:1.19; 95% CI: 1.10-1.48). However, there was no independent asociation in the middle-aged (≤ 60 years old) patients (*P* > 0.05). Regardless of whether it is male or female, this relationship was still statistically significant before and after multivariate adjustment. after multivariate adjustment, the association between the TyG index of female and HFpEF was (OR:1.21; 95% CI: 1.10-1.34), which was higher than that of male (OR:1.11; 95% CI: 1.02-1.21). Multivariate logistic regression analysis showed that the TyG index levels for T2 and T3 were associated with an increased OR for HFpEF when T1 was used as a reference, with the highest association observed for T3 in both female (OR: 1.55; 95% CI: 1.27-2.88) and male (OR: 1.21; 95% CI: 1.10-1.34).

**Table 3.**
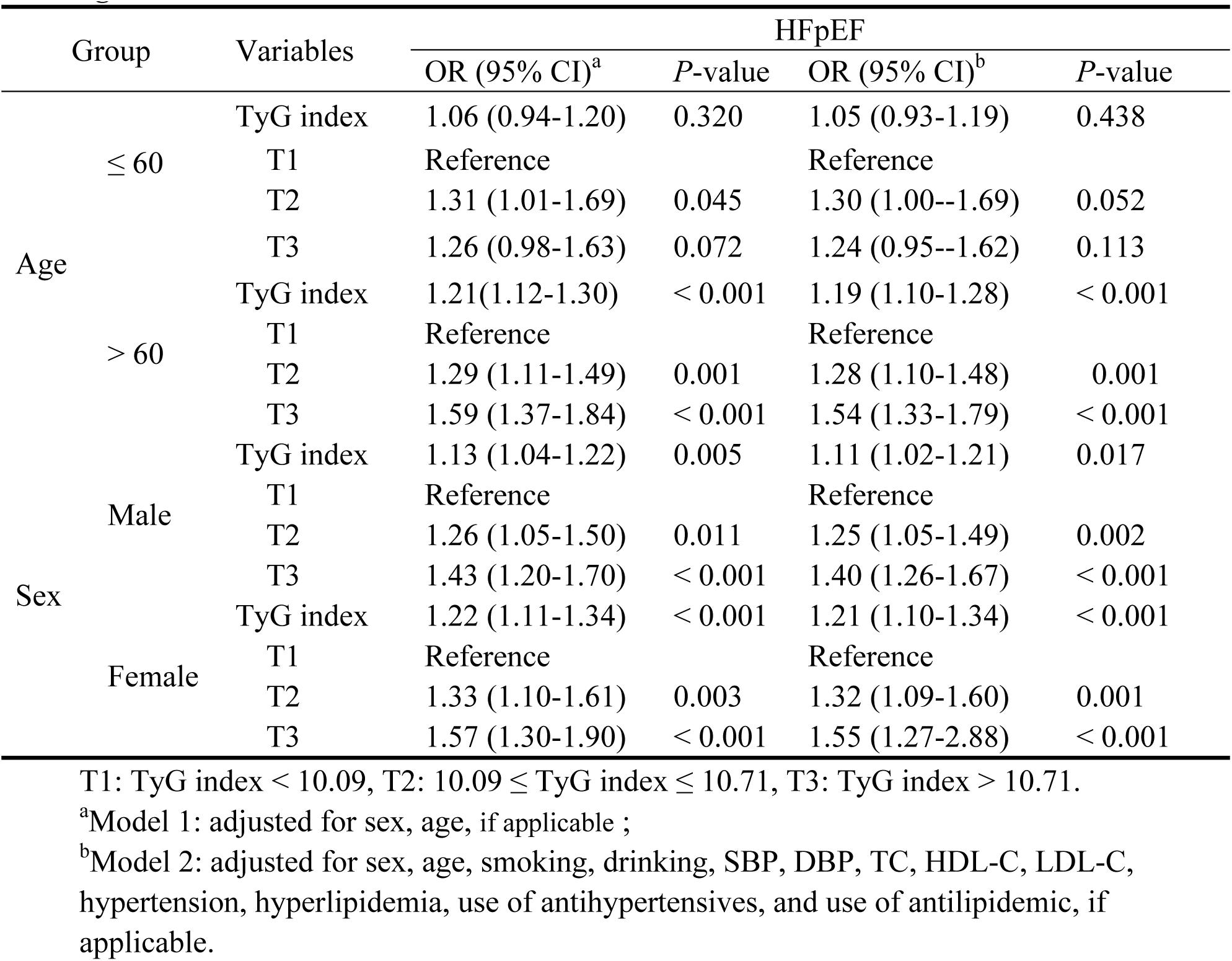
Association between the TyG index and the risk of HFpEF according to age and sex.

### Association between TyG index and the risk of carotid artery plaques according to metabolic status

As shown in **Table 4**, The association between the TyG index and HFpEF clearly varied in different blood pressure and lipid statesa. After multivariate adjustment, the TyG index of CHD patients was significantly associated with HFpEF in hypertension (OR:1.17; 95% CI: 1.10-1.25) and dyslipidemia (OR:1.16; 95% CI: 1.08-1.23). For both hypertension and dyslipidemia, using T1 as the reference, T2 and T3 were significantly related to the increased risks of HFpEF, and T3 observed the highest association in both hypertension (OR: 1.49; 95% CI: 1.30-1.70) and dyslipidemia (OR: 1.48; 95% CI: 1.29-1.68). Even after multivariate adjustment, this relationship remained significant.

**Table 4.**
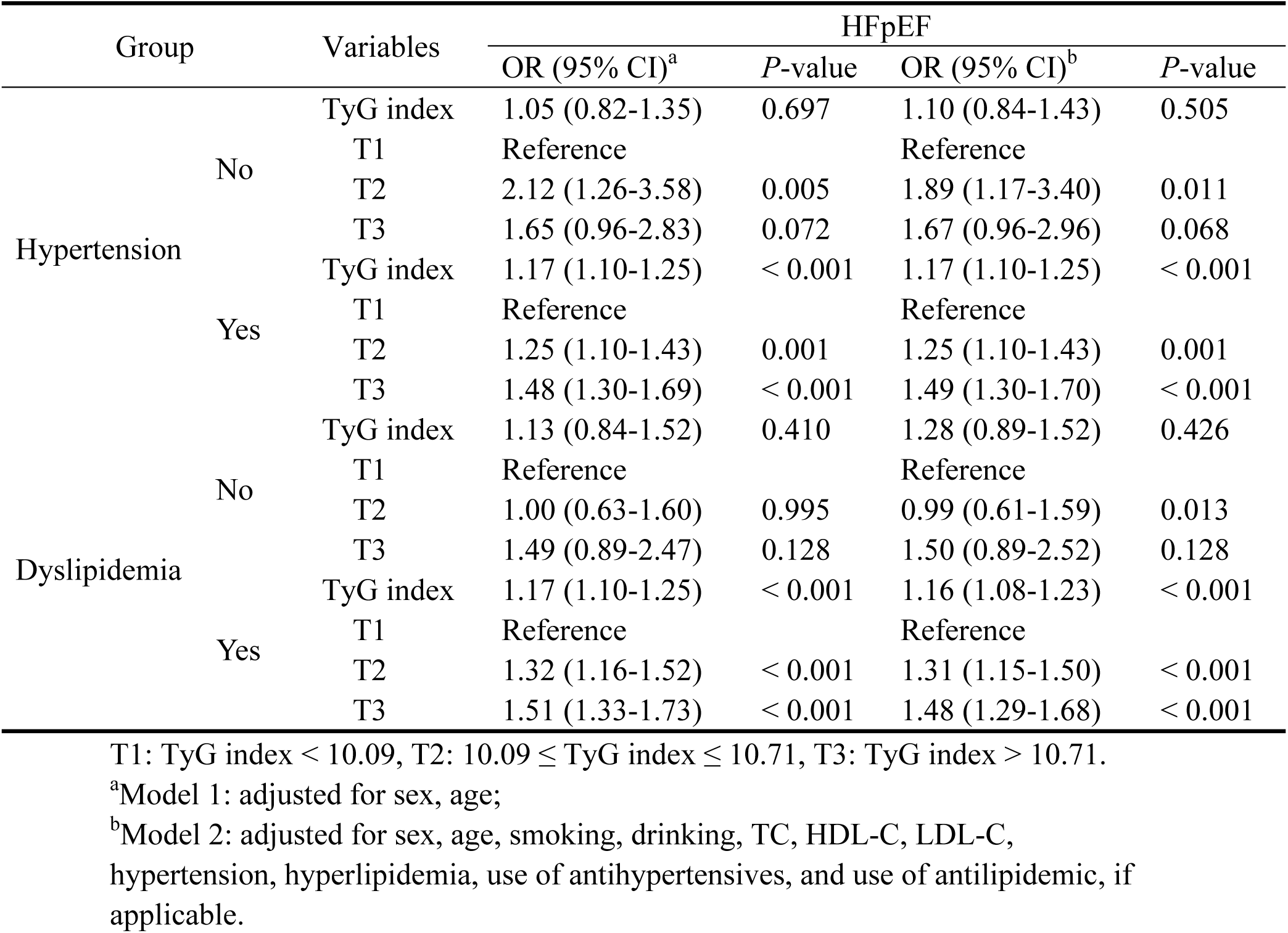
Association between the TyG index and the risk of HFpEF according to hypertension and dyslipidemia.

### Association between TyG index and the risk of carotid artery plaques according to age and metabolic status

As shown in **Table 5**, The TyG index and HFpEF do not show a significant association according to different blood pressure and lipid statesa in aged ≤ 60 years old CHD patients. Then, After multivariate adjustment, the TyG index of CHD patients was significantly associated with HFpEF aged > 60 years old in hypertension (OR:1.20; 95% CI: 1.11-1.30) and dyslipidemia (OR:1.20; 95% CI: 1.11-1.29). For both hypertension and dyslipidemia aged > 60 years old, using T1 as the reference, T2 and T3 were significantly related to the increased risks of HFpEF, and T3 observed the highest association in both hypertension (OR: 1.54; 95% CI: 1.32-1.80) and dyslipidemia (OR: 1.56; 95% CI: 1.34-1.82). Even after multivariate adjustment, this relationship remained significant.

**Table 5.**
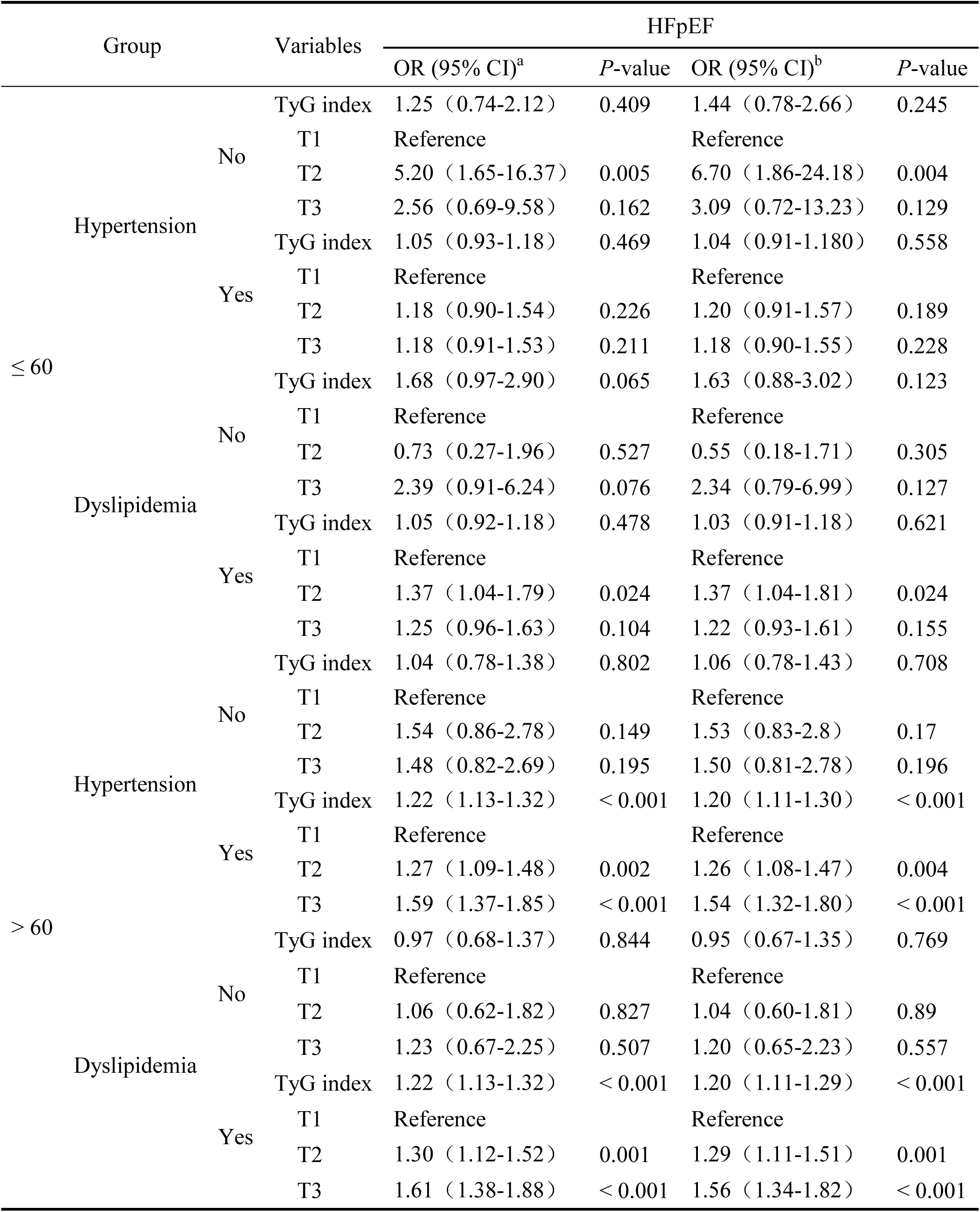
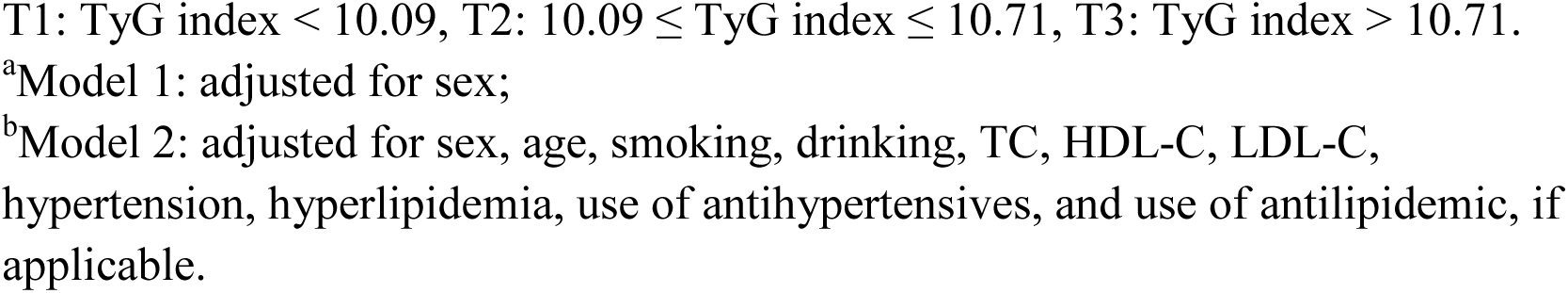
Association between the TyG index and the risk of HFpEF according to age and metabolic status.

### Association between TyG index and the risk of carotid artery plaques according to sex and metabolic status

As shown in **Table 6**, The association between the TyG index and HFpEF was similar in CHD patients of different sex under different blood pressure and lipid status. After multivariate adjustment, the TyG index of male CHD patients was significantly associated with HFpEF in hypertension (OR:1.11; 95% CI: 1.02-1.22) and dyslipidemia (OR:1.32; 95% CI: 1.10-1.58). And the TyG index had the same association with HFpEF in hypertension and dyslipidemia among female CHD patients (OR:1.21; 95% CI: 1.09-1.34). After multivariate adjustment, using T1 as the reference, T2 and T3 were significantly related to the increased risks of HFpEF, and T3 observed the highest association. Among male CHD patients. the association between the TyG index and HFpEF in dyslipidemia (OR: 1.45; 95% CI: 1.21-1.73) was higher than those and hypertension (OR: 1.36; 95% CI: 1.13-1.62) among male CHD patients. However, hypertension (OR: 1.55; 95% CI: 1.27-1.89) was higher than hyperlipidemia (OR: 1.51; 95% CI: 1.23-1.85) in female with CHD patients.

**Table 6.**
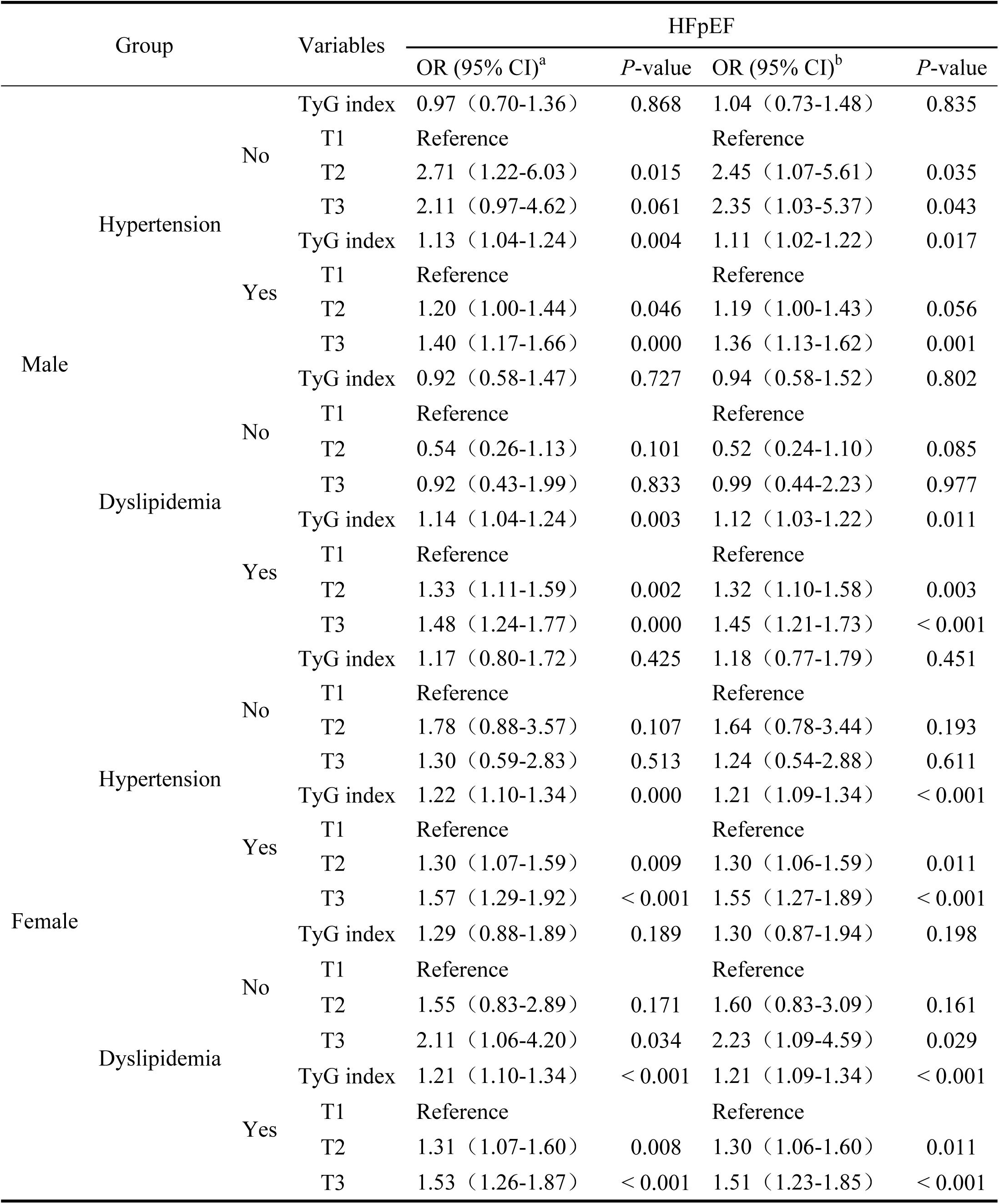
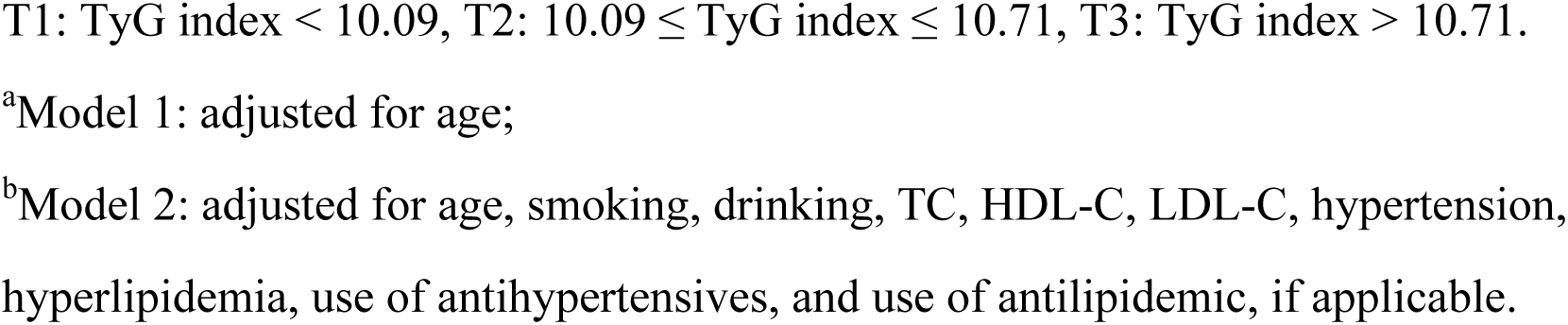
Association between the TyG index and the risk of HFpEF according to sex and metabolic status.

## Discussion

This is the first large-scale study to demonstrate this relationship between the TyG index and HFpEF in CHD patients, and assessed this relationship according to sex, ages and metabolism states (blood pressure, and blood lipid).

HF is a global epidemic with rising prevalence and the prognosis for patients with HF remains poor. Studies have shown that HF is the leading cause of hospitalization in adults, with a 1-year mortality rate of 10-35%, and mortality is much higher in patients with advanced HF [24]. This underscores the importance of primary and secondary prevention of underlying HF conditions, including ischemic HF, management of HFpEF, and HF in the elderly [25]. In recent years, studies have found that the number of HFpEF patients has continued to increase. On the one hand, due to insufficient popularization of primary prevention, the number of high-risk groups of HFpEF has increased. On the other hand, due to the continuous improvement of HFrEF treatment methods, the LVEF of patients has gradually improved, more than 50%. Epidemiological data show that patients with HFpEF have similar all-cause mortality and HF hospitalization rates as HFrEF. Patients with HFpEF are more common in elder, female and hypertensive patients [26–28]. Consistent with the above studies, our study showed that HFpEF had the highest proportion of patients with HF, and that HFpEF patients were older, more male, and had higher systolic blood pressure.

Studies in recent years have shown the close relationship between the TyG index and the homeostasis model assessment of insulin resistance (HOMA-IR). And the predictive value of the TyG index for IR was better than that for HOMA-IR [29]. Studies have shown that the TyG index is positively correlated with the prognosis of HF [30]. And the TyG index is a new biomarker of myocardial fibrosis in patients with HF, which can be regarded as a useful risk stratification indicator in HF management [31]. Several mechanisms implicate the interaction between insulin resistance (IR), myocardial fibrosis, and poor prognosis in HF. Firstly, IR is associated with low-grade inflammation and plays an important role in the pathogenesis of cardiomyocyte apoptosis and myocardial fibrosis. Second, IR is associated with increased activity of the sympathetic nervous system and the renin-angiotensin-aldosterone system; both of them are associated with myocardial fibrosis and cardiac dysfunction. Third, IR is associated with extracellular matrix deposition and intramyocardial lipid deposition, leading to subsequent myocardial fibrosis [31–34]. These studies provide some evidence for our study that TyG has an independent correlation with HF in CHD patients, and there is a certain correlation with different types of HFpEF, therefore, we propose that the TyG index can be regarded as a more convenient marker of IR and considered a useful predictor of HFpEF.

Established risk factors for ASCVD include age, male gender, family history of ASCVD, obesity, hypertension, hypercholesterolemia, and DM [35–36]. Therefore, the association of the TyG index and HFpEF under different risk factor stratification needs to be further explored. In this study, the results show that the TyG index was independently associated with HFpEF in hypertension, dyslipidemia, and elder patients (> 60 years old). In addition, this relationship existed in both male and female patients, with the association being higher in females than in males. A Shanghai-based community-based study on the relationship between macrovascular and microvascular injury and the TyG index in the elderly showed that elevated the TyG index was significantly associated with higher arterial stiffness and risk of renal microvascular injury [37]. In the middle-aged and elderly populations, an increase in the TyG index was significantly associated with hypertension and isolated systolic hypertension [38]. The TyG index may represent a cost-effective and informative screening tool for metabolically obese normal weight (elevated blood pressure, hypertriglyceridemia, hyperglycemia, and low HDL cholesterol) [39]. High TyG index was independently associated with subclinical atherosclerosis (SA) in non-diabetic women, but not in non-diabetic male. TyG index was not associated with the presence of SA in DM patients [40]. While the prevalence of Coronary microvascular dysfunction among men and women with HFpEF is similar, the drivers of microvascular dysfunction may differ by sex. The current inflammatory paradigm of CMD in HFpEF potentially predominates in men, while derangement in ventricular remodelling and fibrosis may play a more important role in women [41].These studies provide some evidence for the findings of this study.

To sum up, with an increasing number of studies on the influence of the TyG index on patients with cardiovascular diseases, the clinical significance of the TyG index is becoming increasingly clear. Evaluation of the TyG index may have important clinical significance for risk stratification and individualized treatment of CHD patients.

## Strengths and limitations

Some limitations of the present study should be noted. First, this study was a multi-center study, thus, there may be bias in the measurement methods at different research centers. However, the practitioners conduted external quality assessments between clinical laboratories in each center. Second, the retrospective design of the current research would cause recall bias, and residual confounders could not be totally avoided. Therefore, any changes in the TyG index that may occur after HF treatment are unknown and require further exploration. And the exact mechanism of the relationship between the TyG index and HFpEF requires further prospective large-scale research.

## Conclusion

This study demonstrated a significant association of the TyG index and HFpEF in CHD patients. In this study, the results show that the TyG index was independently associated with HFpEF in hypertension, dyslipidemia, and elder patients (> 60 years old). In addition, the association between the TyG index and HFpEF in CHD patients was higher in female. The results of this study may emphasize the need for a risk management strategy for specific sex, age and metabolic status to prevent the occurrence of HFpEF in CHD patient.

## Authors’ contributions

CQY, RRY, WC, XF and ZL participated in the study design and statistical analysis; ZL and YJL analyzed the data together and drafted the manuscript; LY, YYH, LL, LY, and SG participated in data collection. All authors have read and approved the final manuscript.

## Data Availability

The datasets used and/or analyzed in the current study are available from the corresponding author upon reasonable request.

## Abbreviations

TyG: Triglyceride glucose
HF: Heart failure
HFrEF: Heart failure with reduced ejection fraction
HFmrEF: Heart failure with mid-range ejection fraction
HFpEF: Heart failure with preserved ejection fraction
LVEF: Left ventricle ejection fraction
CHD: Coronary heart disease
DM: Diabetes mellitus
NCD: Non-communicable diseases
T2DM: Type 2 diabetes
CAD: Coronary artery disease
Pre-DM: Prediabetes
AS: Atherosclerosis
SBP: Systolic blood pressure
DBP: Diastolic blood pressure
FPG: Fasting plasma glucose
TC: Total cholesterol
HDL-C: High-density lipoprotein cholesterol
TG: Triglycerides
LDL-C: Low-density lipoprotein cholesterol
CRP: C-reactionprotein
HbA1c: Glycated haemoglobin
OR: Odds ratios
CIs: Confidence intervals
HOMA-IR: Homeostasis model assessment of insulin resistance
SA: Subclinical atherosclerosis

## Acknowledgments

We thank all the participants in the study and the members of the survey teams, and the groups providing financial support.

## Ethics approval and consent to participate

This study was approved by the ethics committee of Tianjin University of Traditional Chinese Medicine (TJUTCM-EC20190008) and registered in the Chinese Clinical Trial Registry (ChiCTR-1900024535) and in Clinical Trials.gov (NCT04026724).

## Adittional file

**Table S1.** General characteristics of the study participants according to HFpEF.

**Table S2.** Association between the TyG index and the risk of different types of heart failure.

